# Social Determinants of Human Papillomavirus Vaccine Uptake Among Adolescent Girls in Low-Middle-Income Countries: A Systematic Review & Meta-Analysis

**DOI:** 10.1101/2025.08.21.25334143

**Authors:** Pawan Kumar, Arindam Ray, Rhythm Hora, Amrita Kumari, Kapil Singh, Rashmi Mehra, Amanjot Kaur, Shyam Kumar Singh, Seema Singh Koshal, Vivek Kumar Singh, Abida Sultana, Syed F Quadri, Arup Deb Roy

## Abstract

**Introduction:** Cervical cancer remains the fourth most common cancer among women globally, despite being preventable with the human papillomavirus (HPV) vaccine. However, HPV vaccine uptake remains a challenge in low- and middle-income countries (LMICs), where cervical cancer elimination faces significant delays. This study aims to identify the social determinants impacting HPV vaccine uptake in LMICs.

**Methods:** This systematic review and meta-analysis included studies published between 2010 and 2025, identified through PubMed, Google Scholar, and ScienceDirect. Eligible studies reported HPV vaccine uptake (initiation, completion, or both) among adolescent girls aged 9–19 and examined at least one individual- or household-level social determinant. Data were thematically synthesized, and a meta-analysis was conducted using the random-effects model, with results expressed as odds ratios (ORs), with 95 % confidence intervals (CIs).

**Results:** Eight studies, conducted in Ethiopia, Tanzania, and Uganda, were included. Key determinants assessed included age, religion, residence, parental education, occupation, wealth index, marital status, and household factors. Meta-analyses revealed wealth index (OR = 1.34; 95% CI: 1.05–1.70; P = 0.02) and parental marital status (OR = 0.86; 95% CI: 0.78–0.95; P < 0.01) as significant predictors of HPV vaccine uptake among adolescent girls in LMICs. Other factors, such as age, residence, parental education, etc., showed inconsistent effects or no significant association, with high heterogeneity across studies limiting the generalizability of some findings.

**Conclusions:** This review highlights the complex, context-specific individual and household factors influencing HPV vaccine uptake among adolescent girls in LMICs. While wealth index and parental marital status showed consistent associations, other factors varied across studies. Community-based, culturally sensitive, tailored interventions are critical to improve the vaccine uptake. Continued research with standardized mixed-methods is vital to address multilevel factors and ensure equitable HPV vaccine uptake in LMICs.

## 1. Introduction

Human papillomaviruses (HPVs) represent a large and diverse group of viruses with more than 200 types known to date. Of these, more than 40 variants affect the anogenital region, of which approximately 12 high-risk types are oncogenic (1). Among them, HPV variants 16 and 18 have been identified as the primary causative agents leading to cervical cancer (2,3). Although most HPV infections are asymptomatic and resolve without complications, persistent infections with high-risk HPV strains can lead to various cancers, including genital warts, and cancers of the cervix, vulva, vagina, anus, penis, and mouth/throat (4).

Cervical cancer, in particular, remains one of the most prevalent cancers among women, with around 660,000 new cases recorded globally, ranking as the fourth most common cancer worldwide (5–7). In 2022, approximately 94% of the 350, 000 deaths reported due to cervical cancer were reported in low- and middle-income countries (LMICs) (6). This stark disparity is due to the limited resource availability, inadequate public awareness, and late-stage diagnosis, resulting in aggravated complications in cervical cancer mitigation in these regions (8,9).

Although preventable measures such as screening tests and HPV vaccines are available for the prevention of cervical cancer, there is limited utilization in most LMICs. However, vaccination is sought as the primary and most effective strategy for the prevention of cervical cancer and other HPV-associated conditions (10,11). Currently, six HPV vaccines have been licensed for use, with four having a World Health Organization (WHO) pre-qualification. Of the six vaccines, three are bivalent, two are quadrivalent, and one is a nonavalent vaccine (1,11,12). All these vaccines have high efficacy in preventing infection with the virus types 16 and 18, which are together responsible for approximately 77% of cervical cancer cases, while the quadrivalent vaccine additionally prevents against anogenital warts (13). On the other hand, the nonavalent vaccine offers additional protection against other HPV strains too (1,11,12).

While cervical cancer is a largely vaccine-preventable disease, uptake of the HPV vaccine remains a challenge in LMICs, where cervical cancer elimination faces significant delays (11,14). As of February 2025, the HPV vaccine has been introduced into the national immunization programs of approximately 52 LMICs. However, vaccine coverage remains low, with only 13% of girls aged 9–14 years in these settings having received the first dose, which is far below the World Health Organization’s (WHO) global target of 90% coverage among 15-year-old girls to eliminate cervical cancer as a public health problem by 2030. (15). The substantial gap in coverage in LMICs is attributed to a complex interplay of barriers, many rooted in social determinants of health (SDH), including socioeconomic status, gender norms, cultural beliefs, healthcare access, and geographic disparities (16–18), all of which shape awareness, attitudes, and the ability to avail vaccination services (19,20). For instance, lower household income or limited parental education is often associated with reduced knowledge of HPV and the vaccine, contributing to hesitancy or a lack of demand. Cultural or religious norms, particularly those related to adolescent sexual health, may also affect perceived acceptability (21,22). Geographic factors such as rural residence can impede access to vaccination due to a lack of nearby health facilities or outreach programs (23).

While systemic barriers such as uneven vaccine distribution, high costs, logistical constraints, and understaffed health systems have been well documented (12), individual- and household-level variables, such as wealth index, maternal education, parental occupation, and household composition, also play a critical role. For instance, adolescents from more advantaged households are more likely to be vaccinated (24,25). Addressing these inequities is critical to achieving global elimination goals.

Given these challenges, it is crucial to identify and understand the social determinants influencing the HPV vaccine uptake in LMICs (16,26). Therefore, this systematic review and meta-analysis aims to examine these determinants, with a particular focus on individual and household-level factors influencing HPV vaccine uptake among adolescent girls in LMICs. By understanding these intersecting determinants, this study seeks to contribute valuable insights to develop equity-focused interventions to improve HPV vaccine uptake in LMICs.

## 2. Methodology

### 2.1. Definition of Lower Middle-Income Countries (LMICs)

As per the World Bank’s 2024 fiscal year classification, LMICs are defined as those with a Gross National Income (GNI) per capita between $1,136 and $4,465, calculated using the World Bank Atlas method in current US dollars (27).

### 2.2. Data Sources and Search Strategy

This systematic review and meta-analysis followed the Preferred Reporting Items for Systematic Review and Meta-Analysis (PRISMA) 2020 guidelines (28). The PRISMA checklist was followed (Supplementary File 1), and the study selection process is shown in a PRISMA flow diagram. The protocol was not registered due to resource constraints, but PRISMA guidelines ensured methodological rigor (28).

A comprehensive search of three electronic databases - PubMed, Google Scholar, and Science Direct was performed from January 2010 to February 2025 using keywords. The search strings or terms stemmed from the following keywords: “HPV” or “human papillomavirus” AND “vaccine” OR “immunization” AND “uptake” OR “coverage” AND “factors”, OR “determinants” OR “barriers” AND “LMICs” OR “low-middle-income countries” **(Supplementary File 2).** Medical Subject Headings (MeSH) and Boolean operators (“AND,” “OR”) were used to ensure comprehensive coverage. The search strategy was informed by previous systematic reviews conducted on similar topics in European countries, East Africa, and the Sub-Saharan Africa region (20,29,30). The term “LMIC” was purposefully used to streamline selection, though it may have excluded studies referencing specific countries. Yet, it aimed to balance comprehensiveness with feasibility, ensuring that the approach remained practical while still capturing a broad scope of relevant literature. Moreover, the search was limited to English-language articles, as English dominates HPV vaccine research in LMICs, and non-English studies likely offer minimal unique data due to overlap in global databases (31,32). Finally, the reference lists of the selected studies were reviewed to identify additional references not identified via the database search.

### 2.3. Eligibility Criteria

We selected studies reporting HPV vaccine uptake (either initiation, completion, or both) in adolescent girls aged 9-19 years and reporting at least one social determinant, either at the individual or household levels, associated with vaccine uptake. Studies could include primary or secondary data analysis. Only studies in the last 15 years, published between January 2010 and February 2025, were included. All studies aimed at identifying and assessing factors associated with HPV vaccine uptake were included. No LMIC was excluded. Only studies reported in English were considered. Exclusively qualitative studies, review papers, case series, case reports, conference proceedings, abstracts, and studies that did not report the outcome of interest were excluded. Studies focusing on knowledge, attitudes, or intentions to receive the HPV vaccine were also not eligible. Publications reporting the same cohorts were only included if the variables studied were different.

### 2.4. Study Selection

The study selection process was carried out by two authors (RH, RM). Any disagreements were resolved through iterative discussions until an agreement was reached. Eligible articles were selected by first screening titles and abstracts, followed by reviewing full texts and checking the references of these full-text articles. The study selection took place between January 2025 and February 2025, with any disagreements addressed through discussion with a third reviewer (AR) to reach consensus.

### 2.5. Data Extraction

All relevant data were organized and extracted into a PRISMA predefined form (28). Descriptive data such as study identification (first author, year of publication), study location, study setting, study time period, study design, and study population characteristics (such as sample size, age) and relevant study outcomes were extracted for each study. The significance of the associations between these factors and HPV vaccine uptake was evaluated using adjusted odds ratios (AORs)/adjusted prevalence ratios (APRs) with 95% confidence intervals (CIs), considering that most of the included studies in the present review reported AORs or APRs. Factors showing significant associations (p≤0.05) are marked with an asterisk. Factors without significant associations were also extracted for comparison across studies. One author (RH) organized and presented the data narratively in a table, while a second author (RM) verified the data for completeness and accuracy. Any discrepancies were addressed through discussions between both authors (RH & RM).

### 2.6. Data Analysis

The data extracted from each study were presented using a narrative synthesis of the included studies. A summary table was prepared to describe the characteristics (author, year, country, study design, sample size, uptake of HPV vaccine) of the included studies. Socio-demographic and household determinants were identified and selected based on variables commonly reported in previous reviews conducted on a similar topic (20,30) to ensure consistency and allow for meaningful comparisons across different studies.

Meta-analyses were performed using Review Manager 5.4 software (The Cochrane Collaboration, Copenhagen, Denmark). Given the heterogeneity in study populations and designs across the included studies, a random effects model was employed to calculate the pooled ORs for HPV vaccine uptake among adolescent girls in LMICs and associated factors. In the meta-analysis, APRs reported by any included study were converted to odds ratios, provided there was sufficient information for the conversion. If conversion was not possible, APRs were pooled with the AORs. Meta-analyses were not conducted for factors reported by fewer than two studies with comparable classifications.

### 2.7. Quality Assessment

The quality of all included articles was assessed by two independent researchers (RH and RM). The Joanna Briggs Institute (JBI) Critical Appraisal Checklist for Analytical Cross-Sectional Studies was used to evaluate the risk of bias (33). This checklist consists of 8 questions, with each answered by assigning a score of 1 (yes), 0 (no/unclear/not applicable). The overall score for each article was calculated as a percentage (%age), and the risk of bias was categorized based on these scores: a *high risk of bias*, if 20-50% of items scored “yes,” *moderate risk*, if 50-80% scored “yes,” and *low risk of bias*, if 80-100% scored “yes,” as outlined in the JBI checklist, as shown in **Supplementary Table 1**. Out of the 8 studies, five had an overall low risk of bias and three had a moderate risk of bias (**Table 1**). No study was excluded based on the risk of bias.

**Table 1.**
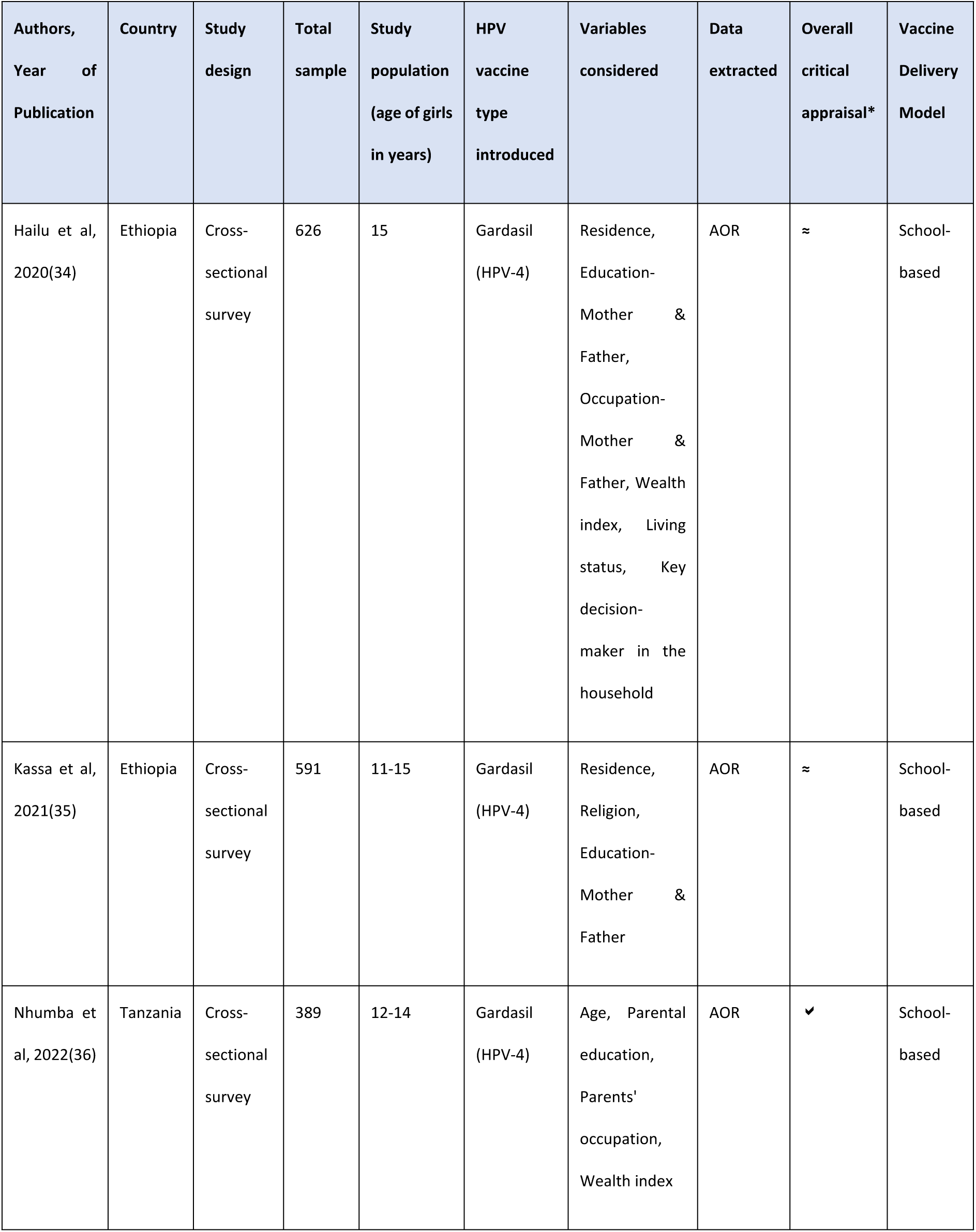
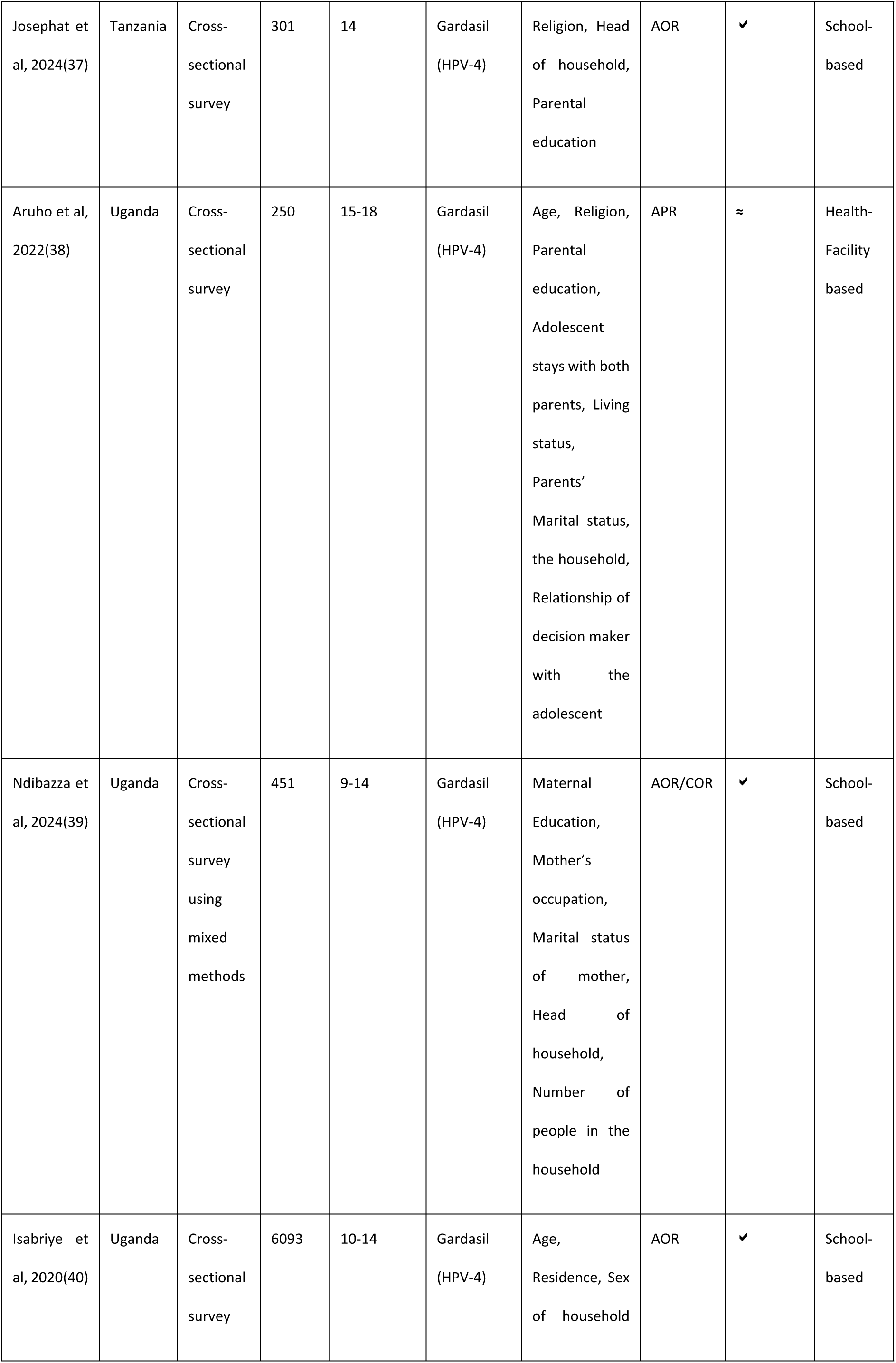
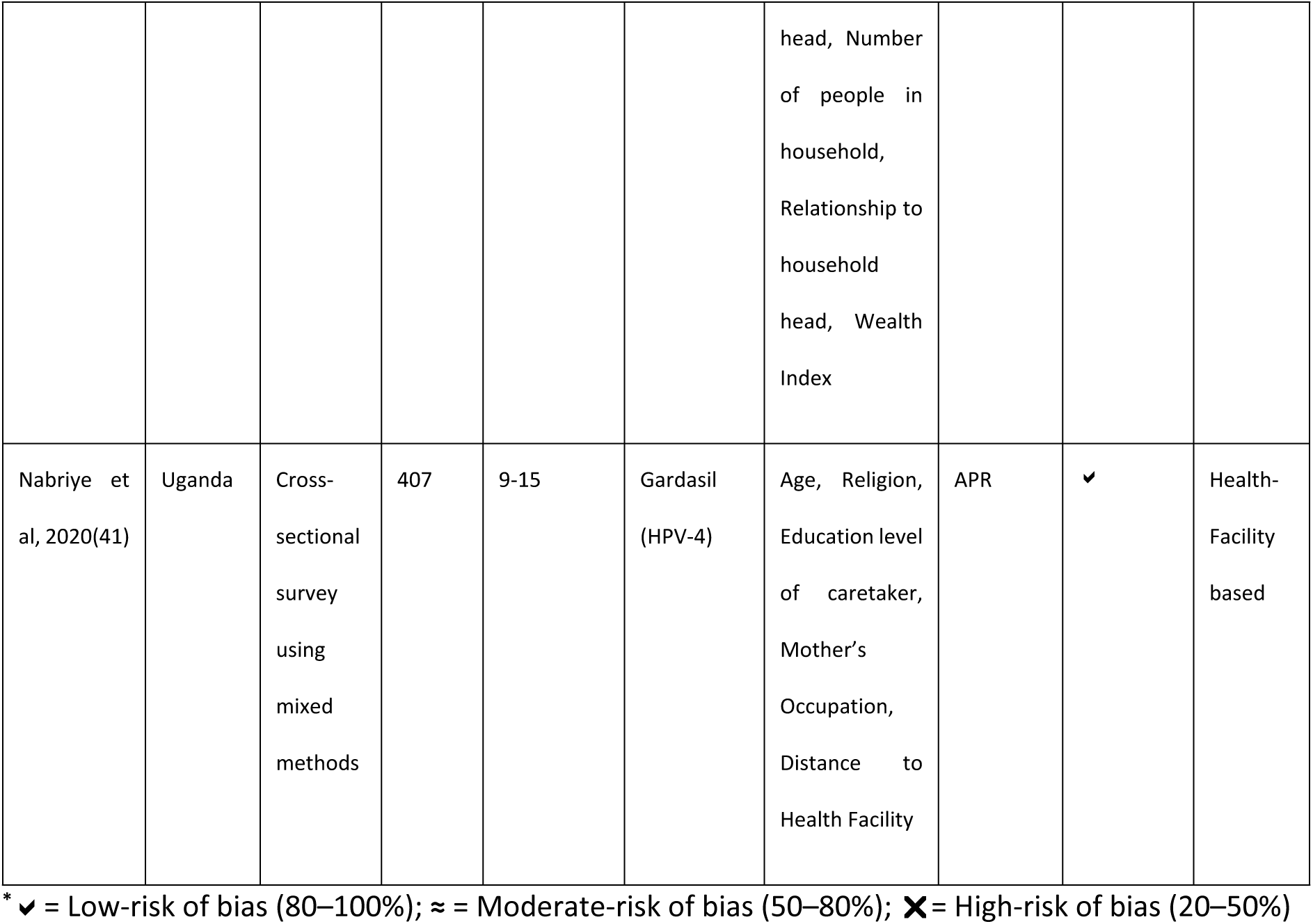
Descriptive characteristics of the studies included in the review.

### 2.7. Ethical Clearance

Ethical clearance was not required for this systematic review article.

## 3. Results

### 3.1. Search Results

A total of 1,052 studies were identified through various databases. Of these, 1,011 were excluded for reasons such as addressing a different research question, not focusing on LMICs, or duplicates (n= 42). The abstracts of the remaining 34 studies were reviewed, and 20 were excluded for the following reasons: duplicates (n= 3), review studies (n= 9), studies addressing a different research question (n= 6), and those not reporting data on factors associated with HPV vaccine uptake (n= 2). Subsequently, 14 full-text studies were reviewed, and their reference lists were checked for additional eligible studies. In total, 8 studies were included in the review—five from the main database search and three from the reference list. All included studies explored the determinants influencing HPV vaccine uptake in LMICs. The study selection process is illustrated in **Figure 1**.

**Figure 1.**
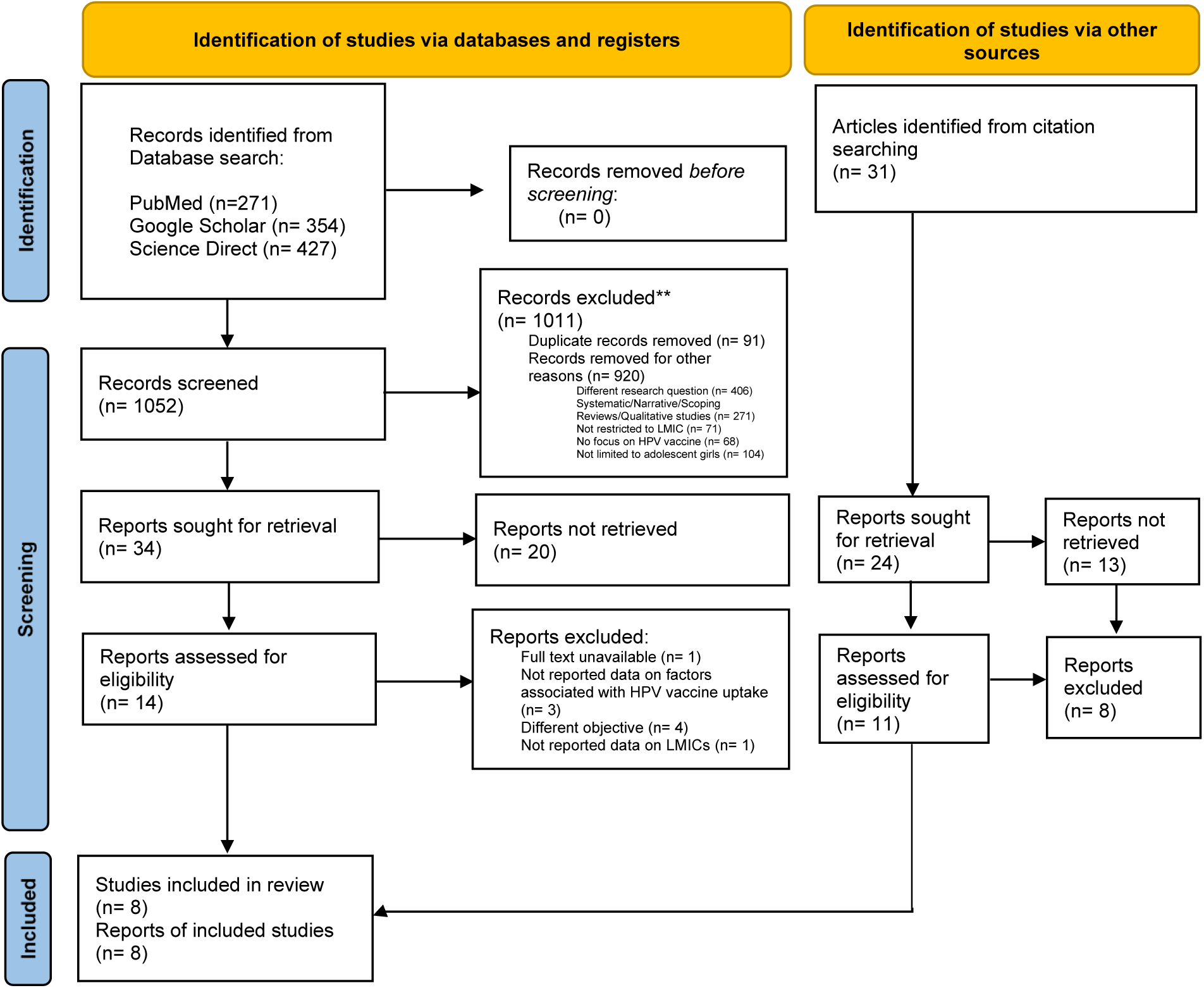
PRISMA 2020 flow diagram for the identification, screening, eligibility, and inclusion of studies.

### 3.2. Descriptive Characteristics of the Eligible Studies

#### 3.2.1. Study Locations

All the included studies were conducted in low- and middle-income countries (LMICs) in East Africa, and no study was found from other countries such as Zambia, Malawi, Zimbabwe, Bolivia, or India, with a reportedly high burden of cervical cancer. The studies included were conducted in Ethiopia, Tanzania, and Uganda. Ethiopia and Tanzania contributed two studies (34–37) while Uganda contributed four studies (38–41) (**Table 1**). Six of the eight studies (34–38,40) included were conducted between April 2016 and December 2022, while 2 studies (39,41) did not report the time. However, all these studies (34–41) were conducted after the introduction of the HPV vaccine in the national immunization program of these countries.

#### 3.2.2. Study Type and Respondents

The majority of the studies (n=6) had a cross-sectional study design (34–38,40), and two studies followed a mixed methods approach (39,41). The study respondents in all studies (34–41) were adolescent girls in the age group of 9-18 years, with sample sizes ranging from 250 to 6,093 (**Table 1**).

### 3.3. Determinants Associated with HPV Vaccine Uptake

Among the included studies (34–41) examining the determinants associated with HPV vaccine uptake, considerable variability was found in the determinants analyzed. Socio-demographic and household determinants, reported in **Supplementary Table 2,** were most commonly assessed across studies and are hereafter summarized under different heads.

#### 3.3.1. Age

Overall, four studies (36,38,40,41) analyzed age as a determinant of HPV vaccine uptake. Three studies (36,40,41) with a cohort of girls aged 9-15 years found a statistically significant association between age and HPV vaccine uptake, with a greater propensity to be vaccinated among older girls than younger ones **(Supplementary Table 2)**. On the contrary, only one study (38) found no statistically significant association between HPV vaccine uptake and age **(Supplementary Table 2)**. In the meta-analysis of these four studies (36,38,40,41), the pooled odds ratio (OR) for the association between age and HPV vaccine uptake was 0.95 (95% CI: 0.54-1.67; P= 0.86), indicating no significant overall effect of age on HPV vaccine uptake (Supplementary Fig. 1a). However, there was substantial heterogeneity observed across the studies (I²= 74%), suggesting that differences in study design, or vaccine delivery models may contribute to the variability in results.

#### 3.3.2. Religion

Four studies (35,37,38,41) assessed religion and HPV vaccine uptake among adolescent girls **(Supplementary Table 2).** Only one study (37) found lower HPV vaccine uptake due to religious prohibitions (AOR=1.19; 95% CI: 0.67-1.79). However, the meta-analysis of these four studies (35,37,38,41) revealed no significant association between religion and HPV vaccine uptake (OR= 1.18; 95% CI= 0.87-1.60; P= 0.29) (Supplementary Fig. 1b). Furthermore, no amount of heterogeneity was found (I^2^= 0%), signaling that the effect sizes across studies were uniform in both magnitude and direction.

#### 3.3.3. Residence

Three studies (34,35,40) assessed the association between the area of residence and the HPV vaccine uptake among adolescent girls. Of these, two studies (34,35) found a statistically significant association **(Supplementary Table 2**), indicating that girls residing in urban areas are more likely to be vaccinated than girls in rural areas. On the other hand, one study found no significant association between the HPV vaccine uptake and area of residence **(Supplementary Table 2)**. The pooled estimates (34,35,40) revealed no significant association (OR= 0.73; 95% CI: 0.16-3.40; P = 0.69) **(Supplementary Fig.1c)**, indicating no overall effect of area of residence on HPV vaccine uptake. However, considerable heterogeneity was observed (I² = 97%), suggesting variability in the results, which may be attributed to differences in population characteristics, study design, or geographical access.

Only one study (41) compared the distance to the health facility and the uptake of the HPV vaccine among adolescent girls **(Supplementary Table 2)**. This study, conducted in Uganda, found that the distance to health facilities was significantly associated with lower uptake (p= 0.038).

#### 3.3.4. Parental Education Status

Of the seven studies (34–39,41) comparing the parental educational status with the HPV vaccine uptake among adolescent girls, only two studies (36,37) found that adolescents whose parents had no formal education had a lesser likelihood of HPV vaccine uptake than those whose parents had secondary or higher education (p < 0.05) **(Supplementary Table 2)**. Additionally, five studies (34,35,38,39,41) found no significant association between the two variables(**Supplementary Table 2**). A meta-analysis of these seven studies(34–39,41) showed no significant association between parental education status and HPV vaccine uptake (OR= 1.13; 95% CI= 0.87-1.47; P= 0.36; I^2^= 29%) (Supplementary Fig. 1d). Moreover, a minimal amount of heterogeneity was found, suggesting consistent effect sizes across the studies.

#### 3.3.5. Parents’ Occupation

Five studies (34–36,39,41) evaluated the association between the occupation of the adolescent girl’s parent with the HPV vaccine uptake, of which three studies (36,39,41) showed a statistically significant association **(Supplementary Table 2)**. Furthermore, two studies (34,35) reported no association between the parents’ occupation and the HPV vaccine uptake **(Supplementary Table 2)**. In the meta-analysis of these five studies (34–36,39,41), the pooled OR was 1.47 (95% CI: 0.74-2.94; P= 0.27), indicating no significant overall effect of parent’s occupation on HPV vaccine uptake (Supplementary Fig. 1e). However, there was substantial heterogeneity observed across the studies (I²=72%), indicating a potential inconsistency in the occupational categories, vaccine delivery models, or population characteristics.

#### 3.3.6. Wealth Index

In the included studies, the wealth index was most commonly constructed using Principal Component Analysis (PCA) based on household assets. Households were then grouped into quintiles (poorest to wealthiest). Some studies also applied PCA at the community level to assess broader socioeconomic influences. Three studies (34,36,40) reported data on the wealth index **(Supplementary Table 2)**. Only one study (40) showed higher odds of HPV vaccination among girls whose families belong to a middle wealth quintile (OR=1.31; 95% CI 1.01–1.69) compared to ones belonging to communities with the poor wealth quintile. On the contrary, two studies (34,36) found no evidence of association between the two variables. A meta-analysis of these three studies found a significant association between the wealth index and HPV vaccine uptake (OR=1.34; 95% CI: 1.05-1.70; P = 0.02) (Fig. 2a). Furthermore, no heterogeneity was found (I^2^= 0%), indicating robust findings.

**Figure 2.**
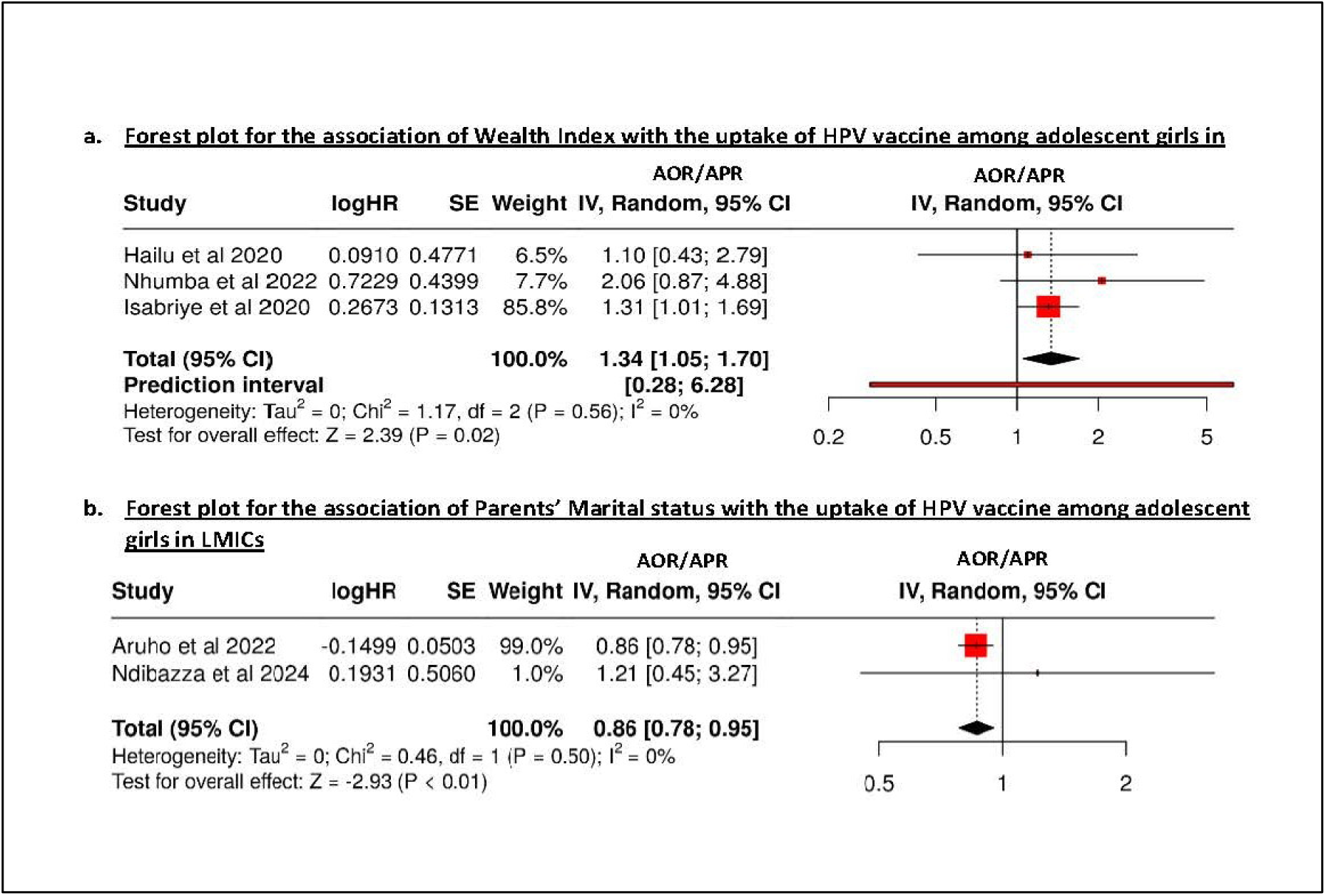
(a, b): Forest plot for the association of: a) Wealth Index, b) Parental Marital status with the HPV vaccine uptake among adolescent girls in LMICs.

#### 3.3.7. Parents’ Marital Status

Overall, two studies (38,39) assessed the association between the marital status of the parents and the HPV vaccine uptake **(Supplementary Table 2)**. Only one of the two studies reported that the uptake was significantly lower among the adolescents whose parents were single (AOR=0.85) (38). (**Supplementary Table 2)**. In the meta-analysis of these two studies (38,39), the pooled OR for the association between parents’ marital status and HPV vaccine uptake was 0.86 (95% CI: 0.78-0.95; P<0.01; I^2^= 0%), indicating a significant overall effect of parental marital status on HPV vaccine uptake (Fig. 2b) with no heterogeneity across the studies.

#### 3.3.8. Household Factors

Two studies (34,38) assessed the living status of adolescent girls with the HPV vaccination uptake. Only one study (38) found that the vaccine uptake was significantly lower (AOR=0.76) among adolescents who stayed only with their mothers instead of both parents **(Supplementary Table 2),** while the other two studies (34,38) (Supplementary Fig. 2a) reported no significant association **(Supplementary Table 2).** Meta-analysis of these two studies (34,38) revealed no significant association for the living status of adolescent girls (OR= 0.93; 95% CI= 0.55-1.58; P= 0.79; I^2^= 56%) **(Supplementary** Fig. 2a), or key decision-maker in the household (OR= 1.09; 95% CI= 0.86-1.38; P= 0.49; I^2^= 0%) **(Supplementary** Fig. 2b).

Furthermore, four studies (37–40) analyzed the association between the household head and HPV vaccine uptake. However, only one study (37) reported significantly higher odds of vaccine uptake among girls living in households headed by relatives than those headed by parents/ grandparents **(Supplementary Table 2)**. A meta-analysis of these four studies (37–40) revealed no significant association between the household head and HPV vaccine uptake (OR= 0.97; 95% CI= 0.79-1.19; P= 0.77) **(Supplementary** Fig. 2c).

Two studies (39,40) also evaluated the number of people in the household. Both (39,40) found lower odds of HPV vaccine uptake among girls living in households with more family members **(Supplementary Table 2)**. However, the meta-analysis showed no significant effect (OR = 0.57; 95% CI: 0.23–1.45; P = 0.24; I² = 68%) **(Supplementary** Fig. 2d).

## 4. Discussion

The present systematic review and meta-analysis identified the social determinants influencing HPV vaccine uptake among adolescent girls in LMICs. A total of eight studies, conducted in Ethiopia, Tanzania, and Uganda, were included in this review, providing insights into how socio-demographic and household factors impact vaccine uptake in these contexts. While several determinants were identified at individual and household levels, there was variability in the strength and consistency of their associations across studies.

**Age** was found to be a consistent determinant in most of the studies, with older adolescent girls showing a higher likelihood of HPV vaccination compared to younger girls. This is consistent with globally observed patterns, where the HPV vaccination uptake was higher among older age groups (42). However, a meta-analysis of the present study found no significant association (OR = 0.95; I² = 74%), which may be attributed to confounding factors such as parental perceptions of the vaccine or differences in vaccine delivery models (e.g., school-based vs. health facility-based) that influence uptake rates at specific ages (42). Further, **religion** emerged as another determinant, but with a less pronounced impact. While individual studies suggest that religious beliefs could pose barriers, the majority of the studies did not find any significant relationship between the two variables. Additionally, meta-analysis of this review found no significant association between the HPV vaccine uptake among adolescent girls and religion (pooled OR =1.18). The findings correlate with other studies by Fisher et al. and Yeganeh et al. that found no strong evidence between religion and vaccination rate (19,43). **Residence**, particularly the distinction between urban and rural areas, was identified as a significant determinant of HPV vaccine uptake, with girls in urban areas being substantially more likely to receive the HPV vaccine compared to their rural counterparts. This is in line with the results of the previous study by Fernandez et al. that found that girls residing in urban areas had a higher likelihood of HPV vaccine schedule initiation/completion (20), often due to limited healthcare resources. However, the area of residence was not a significant determinant in the pooled analysis (OR = 0.73; I² = 97%), in contrast to a study by Addisu et al. that found significantly higher HPV vaccine uptake in urban girls (OR = 4.17; 95% CI = 1.81–9.58) (44). Besides, distance to health facilities was found to be a significant barrier, where girls living farther from healthcare facilities were less likely to receive the HPV vaccine. A previous study conducted by Tsui et al. also highlighted that the distance to vaccination services is a potential barrier to vaccine uptake (45).

The role of parental education in HPV vaccine uptake was also explored, with mixed results. While some studies found that adolescent girls with parents with higher education levels were more likely to get their daughters vaccinated, others found no significant relationship. Also, the meta-analysis of the present study did not find any significant relationship between parental education and uptake of the HPV vaccination among adolescent girls in LMICs. Similar findings were reported in the previous study by Anyaka et al. which found no significant association between parents’ education and daughters’ HPV vaccination uptake (p= 0.056) (46). The variability could be explained by other confounding factors, such as socioeconomic status or community norms, which might outweigh the direct effect of parental education. Additionally, parental occupation also showed inconsistent associations. While some studies linked parental, particularly mothers’ employment with a greater uptake of HPV vaccination, the overall pooled effects were non-significant. This aligns with the previous meta-analysis showing lower uptake among children of unemployed parents (pooled OR = 0.45; 95% CI = 0.28–0.74) (47,48). This may be due to the increased socio-economic status and access to resources and information in families with employed parents. Furthermore, the wealth index showed inconsistent associations with HPV vaccine uptake. While some individual studies reported higher vaccine uptake among girls from middle-income households, others found no such relationship. However, the findings of the present meta-analysis reveal a significant relationship (p= 0.02) between the wealth index and HPV vaccine uptake among adolescent girls in LMICs. Consistent with these findings, prior systematic review by Xu et al. found that adolescent girls from low-income families had lower HPV vaccination rates, while higher income was associated with increased HPV vaccination uptake (25). Similarly, the marital status of parents showed varied effects, with inconsistent associations across studies, underscoring the complexity of household dynamics in influencing vaccine uptake. The meta-analysis found a significant relationship between the marital status of the parents and the HPV vaccine uptake with a pooled estimate of 86%. However, a previous review by Lopez et al. observed no significant difference between marital status and HPV vaccine uptake (49).

Moreover, household factors, including living arrangements and the role of household heads, also played a role in HPV vaccine uptake. For instance, girls living with extended family or non-parental guardians were more likely to receive the vaccine. A previous review by Salleh et al. highlighted that in families where the fathers were the ultimate decision-makers, especially those concerning the children, a major barrier to vaccinating their daughters (50). Conversely, girls living in larger households were found to have lower vaccine uptake, potentially due to competing family priorities, lower economic constraints, or more complex decision-making processes in larger families. However, the meta-analysis of this review found no significant relationship between the HPV vaccine uptake among adolescent girls with these determinants.

Overall, while certain patterns emerged, high heterogeneity across studies reflected differences in study design, population characteristics, and vaccine delivery models, limiting the generalizability of the findings. This variability underscores the importance of context and suggests that social determinants operate within broader structural and cultural frameworks.

## Limitations

This review, however, has some limitations. First, using the term “LMIC” in the search strategy may have led to the omission of studies referencing specific countries, limiting geographic diversity. Listing all LMICs individually could have introduced irrelevant or duplicate results, reducing screening efficiency. Second, the review did not include unpublished gray literature or studies published in languages other than English, which may have limited the inclusion of relevant research/ studies published in other languages, such as French in some African countries or Spanish in parts of Latin America. While English dominates HPV vaccine research and overlaps in global databases reduce the risk of missing key data, this restriction may limit region-specific cultural or programmatic insights. Third, the inclusion of both mixed methods (quantitative element) and quantitative cross-sectional studies in the current review may have contributed to increased heterogeneity in the review findings.

A key limitation of this review is the high heterogeneity across included studies (as indicated by the I² statistic), which may have limited standardization of effect estimates and outcomes, potentially affecting the comparability, robustness, and validity of the meta-analysis results.

## Recommendations & Implications

The findings emphasize the need for context-specific interventions tailored to the diverse challenges faced by LMICs. Programs should address social, cultural, and economic barriers at the community level, particularly in rural and low-income areas, through localized engagement strategies, education campaigns, and partnerships with trusted local actors. Parental involvement in vaccination efforts can further help build trust, counter misinformation, and increase vaccine acceptance.

Among individual-level factors, the wealth index consistently influenced vaccine uptake and should be factored into policy and program design to ensure equitable access. Additionally, the significant association between parental marital status and vaccine uptake points to the importance of considering family dynamics in outreach and communication efforts.

Although factors like age, residence, parental occupation, and education showed mixed or non-significant effects overall, their influence in individual studies suggests a need for targeted interventions considering local demographic and structural contexts. Expanding access through school-based or mobile delivery models may further improve uptake across underserved populations.

Standardized, mixed-method approaches are essential to disentangle these complex dynamics and inform effective, contextualized strategies for improving HPV vaccine uptake in LMICs. While this review focused on individual- and household-level determinants of HPV vaccine uptake, broader systemic issues in the LMICs, such as health infrastructure gaps, logistical constraints, and vaccine delivery models, must also be acknowledged to ensure sustained and equitable HPV vaccine coverage in LMICs. Besides, future research should include non-English studies from regional databases (e.g., Scielo for Latin America, AJOL for Africa) to enhance comprehensiveness and explore diverse LMIC contexts and systemic factors.

## 5. Conclusions

This systematic review and meta-analysis underscore the complex and context-specific nature of HPV vaccine uptake among adolescent girls in LMICs. While no single factor uniformly predicted uptake across all settings, socioeconomic status, particularly household wealth and parental marital status, emerged as more consistent determinants. Other variables such as age, residence, education, and occupation showed mixed associations, underscoring the need for localized, data-driven strategies.

These findings suggest that improving HPV vaccine uptake in LMICs requires a multifaceted, context-sensitive approach. Efforts must go beyond one-size-fits-all strategies by factoring addressing community-specific barriers, improving accessibility, and integrating family dynamics into program design. To strengthen and sustain progress, continued research is needed to fill existing evidence gaps and inform more targeted, effective interventions tailored to local needs.

## Author Contribution Statement

Conceptualization, A.R. and R.H.; Methodology, R.H., R.M., and A.R.; Software, R.H. and RM.; Validation, P.K., A.R., A.K., and A.D.R.; Formal Analysis, R.H., S.K.S.; Investigation, K.S., S.Q., Amanjot Kaur., and R.H.; Resources, R.H., V.K.S. and R.M..; Data Curation, K.S., A.S., S.S.K. and R.H.; Writing – Original Draft Preparation, R.H., V.K.S. and A.R.; Writing – Review & Editing, P.K., S.S.K., A.K., and Amanjot Kaur; Visualization, R.H., and S.K.S.; Supervision, P.K., A.R., S.S.K., and S.Q.; Project Administration, A.D.R.; Funding Acquisition, A.R. and A.D.R. All authors have read and agreed to the published version of the manuscript.

## Data Availability

All data underlying this systematic review, including search strategies (Supplementary File 2), study selection flow (PRISMA flowchart in the main manuscript), and data extraction sheets (Table 1 in the main manuscript and Supplementary Table 2 in the supporting zip folder), are available in the submitted supplementary materials. These will be made publicly accessible upon publication, in accordance with ethical and legal guidelines.

---

**Supplementary Figure 1.**
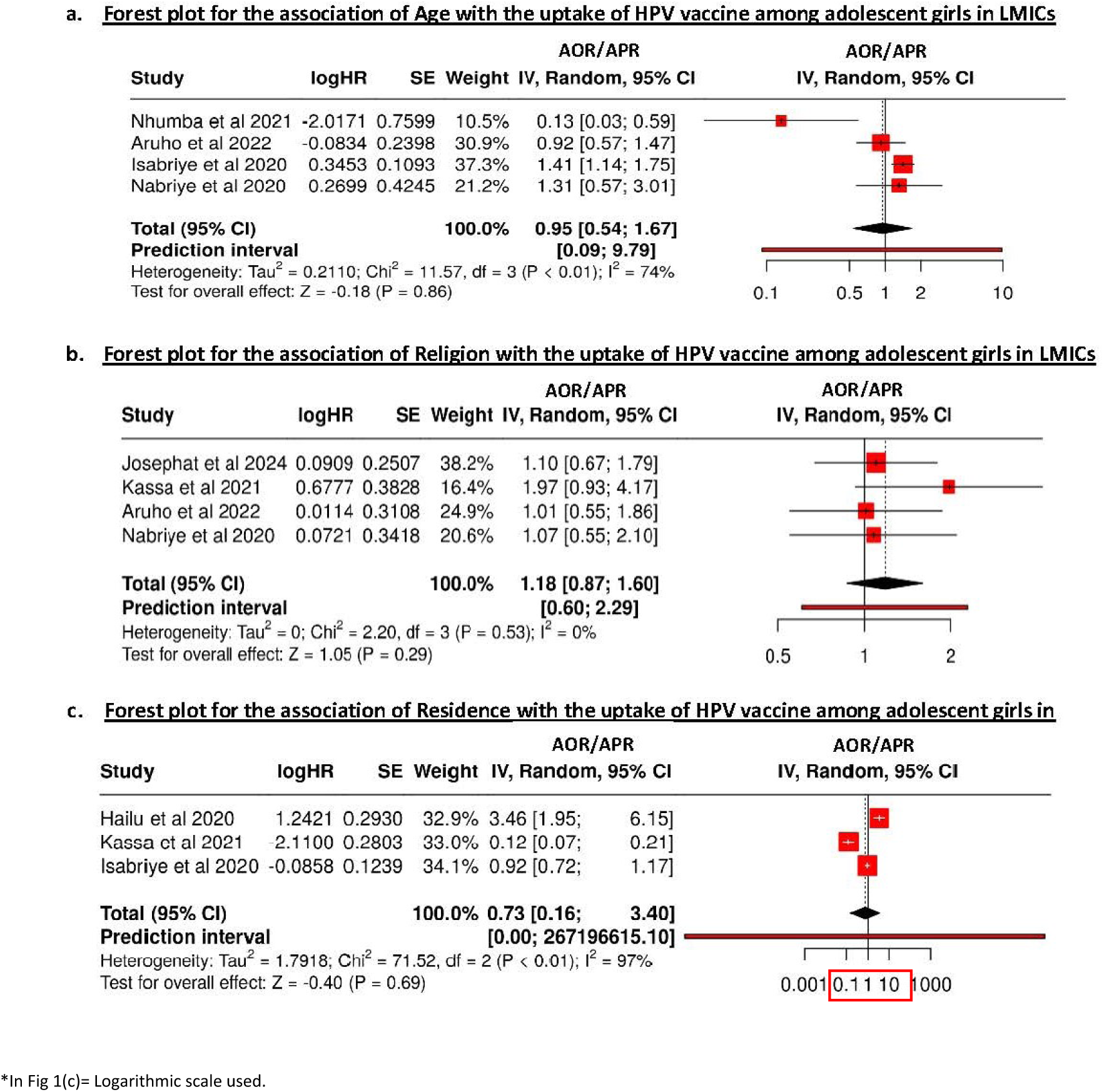
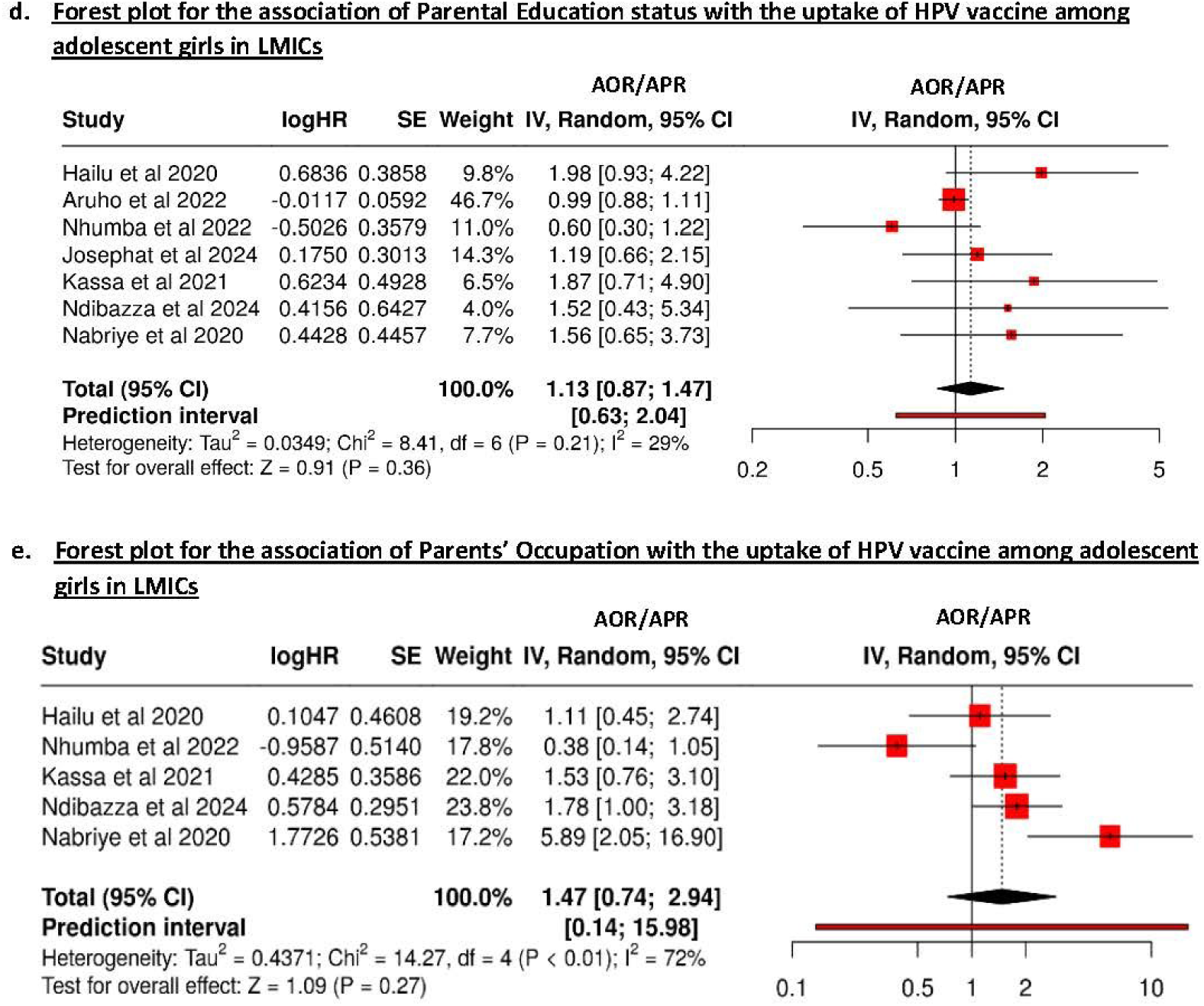
(a, b, c, d & e): Forest plot for the association of a) Age, b) Religion, c) Residence, d) Parental Education, and e) Parental Occupation with the HPV vaccine uptake among adolescent girls in LMICs.

**Supplementary Figure 2.**
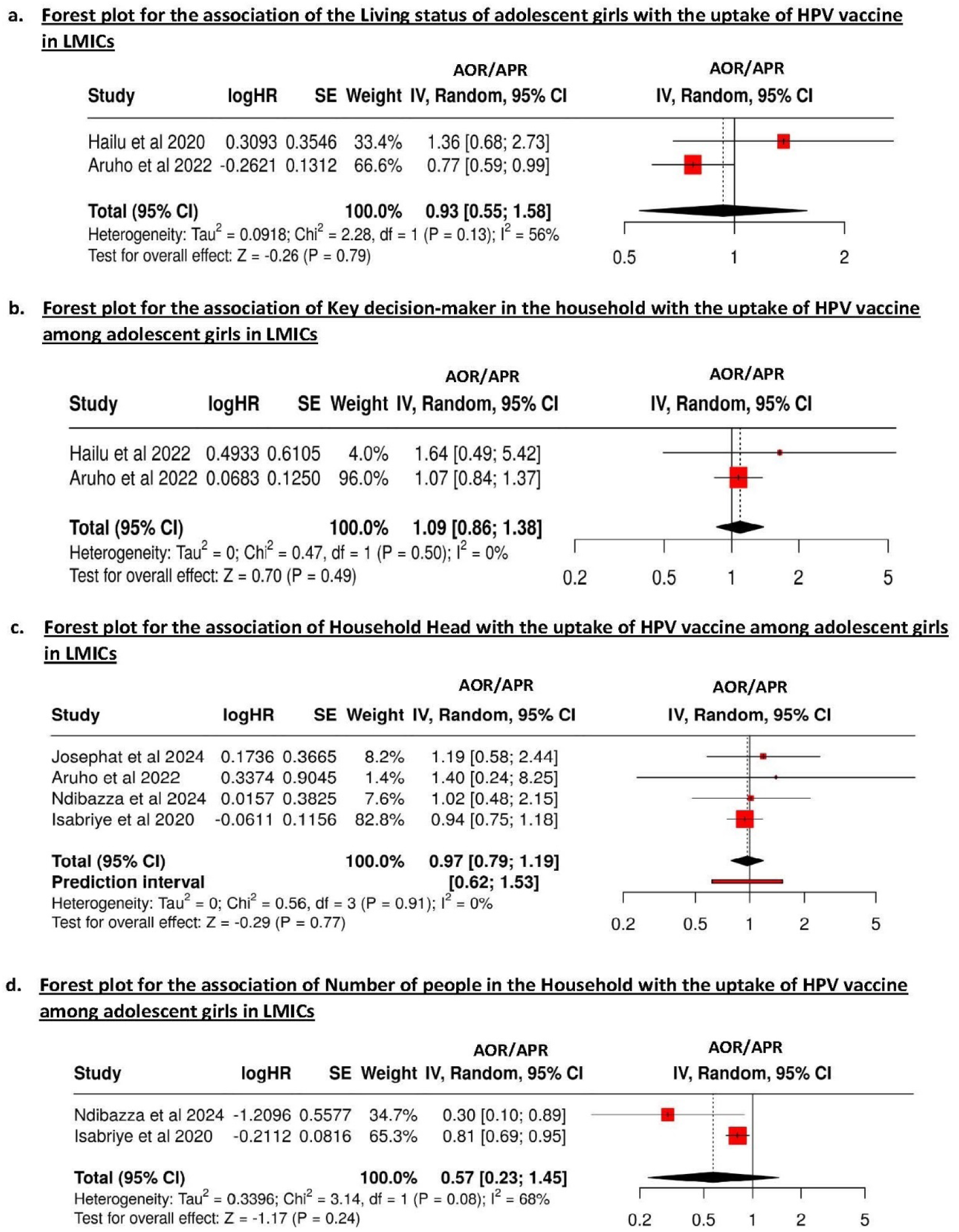
Forest plot for the association of the Household factors with the HPV vaccine uptake among adolescent girls in LMICs.

